# Predicting Impaired Cerebrovascular Reactivity and Hyperperfusion Syndrome with BeamSAT MRI in Carotid Artery Stenosis

**DOI:** 10.1101/2024.01.02.24300743

**Authors:** Yusuke Ikeuchi, Masaaki Kohta, Shunsuke Yamashita, Shunsuke Yamanishi, Yoji Yamaguchi, Jun Tanaka, Kazuhiro Tanaka, Hidehito Kimura, Atsushi Fujita, Kohkichi Hosoda, Eiji Kohmura, Takashi Sasayama

## Abstract

**Background:** We evaluated the impact of collateral flow of the middle cerebral artery (MCA) territory and cross flow visualized with internal carotid artery (ICA)-selective Magnetic resonance angiography (MRA) constructed by pencil beam presaturation (BeamSAT) pulse on preoperative cerebral hemodynamic status and postoperative hyperperfusion syndrome.

**Methods:** Fourty-eight patients who underwent carotid artery stenting or carotid endarterectomy were included. The preoperative status of collateral flow in the MCA territory and crossflow was assessed using ICA-selective MRA. Cerebral blood flow and cerebrovascular reactivity (CVR) were assessed using single-photon emission computed tomography. Potential signs of hyperperfusion syndrome (HPS) were retrospectively assessed by reviewing medical charts.

**Results:** Patients who either demonstrated mismatch in MCA signal intensity (MCA-MRA grade) between ICA-selective MRA by BeamSAT magnetic resonance imaging (MRI) and conventional MRA or whose MRA crossflow was visualized on BeamSAT MRI were placed in the developed collateral (DC) group (n=19). All other patients were placed in the undeveloped collateral (UC) group (n = 29). Preoperative ipsilateral CVR was significantly lower in the DC group than in the UC group (18.0±20.0% versus 48.3±19.5%; P<0.0001). Multivariate logistic regression analysis revealed that the DC group was significantly associated with impaired CVR (odds ratio [OR] = 17.7, [95%confidence interval [CI]: 1.82 to 171]; P = 0.013). The partial area under curves (AUCs) of BeamSAT logisitic model (0.843) were significantly larger than those of the conventional logistic model (0.626) over the range of high sensitivity (0.6-1) (p = 0.04). The incidence of postoperative symptoms suggestive of HPS was significantly higher in the DC group than in the UC group (8/19 vs. 1/29; P=0.001).

**Conclusions:** The differences in the MCA-MRA grade and crossflow between ICA-selective and conventional MRA were associated with impaired CVR. BeamSAT MRI may be a valuable and non-invasive tool for assessing of cerebral hemodynamics and predicting postoperative HPS.

## Introduction

Impaired cerebrovascular reactivity (CVR) is a condition in which the brain’s autoregulatory capacity is compromised, leading to the risk factor of cerebral infarction caused by carotid artery stenosis.^1^ Cerebral hyperperfusion syndrome (HPS) is a complex clinical phenomenon that may occur after revascularization procedures such as carotid endarterectomy (CEA) and carotid artery stenting (CAS).^2^ Preoperative impaired CVR is a risk factor for HPS, necessitating careful postoperative management.^3^ Single-photon emission computed tomography (SPECT) with an acetazolamide (ACZ) challenge can detect impaired CVR with diagnostic accuracy.^4^ Positron emission tomography (PET) is commonly used to assess cerebral misery perfusion (CMP).^5^ However, these methods have drawbacks, as both SPECT and PET involve exposure to radiation and require injection of radioactive tracers.

Considering these issues, we explored the feasibility of assessing impaired CVR using pencil beam presaturation (BeamSAT) magnetic resonance imaging (MRI) technology, which does not require contrast agents or radioactive tracers.^6,7^ BeamSAT MRI is a novel imaging method that uniquely enables the production of selective MRA images of specific arteries, such as the unilateral internal carotid artery (ICA) or vertebral artery (VA).^6,8^ Accordingly, BeamSAT MRI is considered a minimally invasive method that provides an accurate evaluation of intracranial blood flow.

Previous research has reported that the presence of collateral flow is a prerequisite for the impaired CVR caused by carotid artery stenosis.^9,10^ In our earlier study on BeamSAT MRI,^6,7^ we observed discrepancies in the time-of-flight (TOF) signal intensity of the MCA between conventional and ICA-selective MRA in certain cases. Additionally, contralateral ICA-selective MRA and VA-selective MRA showed cross flow or collateral flow to the MCA on the stenotic side in some cases. These observations led us to suspect the presence of collateral flow. Therefore, we hypothesized that patients with these findings were likely to be be more susceptible to impaired CVR.

The aim of this study was to evaluate the possibility that patients with a discrepancy in the MCA intensity between conventional MRA and ICA-selective MRA, or crossflow in contralateral ICA-selective MRA or VA-selective MRA, may have impaired CVR. Additionally, the present study aimed to assess whether patients showing preoperative collateral flow in BeamSAT MRI may be more susceptible to HPS.

## Methods

### Ethics statement

This study was approved by the Institutional Review Board of the Kobe University Hospital (approval number: # 170024), and written informed consent was obtained from all patients. The study was conducted according to the principles of the Declaration of Helsinki regarding experimentation on human participants and the study also complied with national regulations.

### Study design and patients

We performed a prospective cohort study using MRI in patients with carotid stenosis to assess the clinical usefulness of arterial spin labeling (ASL) in cerebral blood flow (CBF) assessments and to investigate the association between MRI plaque findings and the risk of new ischemic lesions after CEA or CAS.^11–13^ Accordingly, we prospectively recruited patients who were under consideration for either CEA or CAS and retrospectively reviewed their BeamSAT results.

Forty-eight patients who underwent preoperative examinations before carotid revascularization between November 2016 and December 2020 were included in this study. The inclusion criteria were patients who underwent conventional MRA, BeamSAT MRI, and SPECT with ACZ before CEA or CAS, symptomatic patients with carotid stenosis of ≥50%, and asymptomatic patients with stenosis of ≥60%, according to the criteria in previous studies.^14,15^ Symptomatic patients were defined as those who experienced amaurosis fugax, transient ischemic attack, or stroke in the territory of the ipsilateral carotid artery within 6 months before study entry. The exclusion criteria were emergency CEA or CAS for stroke-in-evolution or crescendo transient ischemic attack, major disabling stroke, contraindications for MR imaging, or patients in whom waiting for MR imaging would have delayed CEA or CAS. The choice of CEA or CAS was determined by the surgeon or interventionist responsible for the patient. In general, CEA was considered as the first-line therapy for carotid stenosis with marked calcification and vascular kinking or tortuosity. CAS was indicated for patients with high bifurcations, for restenosis after CEA, and for those who preferred CAS to CEA. The physicians and patients discussed treatment policies, and the procedures to be conducted were decided on a patient-by-patient basis.

The following items were retrospectively investigated from the medical charts 1 week postoperatively as symptoms of suspected HPS: multiple complaints of headache and nausea/vomiting, seizures, focal neurological deficits, and intracranial hemorrhage on imaging. All patients underwent ACZ-free SPECT on the day after CEA or CAS and BeamSAT MRI within 2 days after CEA/CAS.

### MR imaging protocol

Conventional and ICA-selective MRA or VA-selective MRA was performed using a 1.5T MRI scanner (Echelon Vega; FUJIFILM Healthcare Corporation, Tokyo, Japan) and an 8-channel head coil, as previously reported.^6^ The 3D TOF-MRA parameters were as follows: repetition time, 23.0 ms; echo time, 6.9 ms; flip angle, 20°; field of view, 230 mm; matrix, 512×200; slice interval, 0.55 mm (after zero-fill interpolation); number of slices, 152; and acquisition time, 4 minutes 50 seconds. For selective 3D TOF-MRA, the BeamSAT pulse was positioned on a TOF source image from conventional MRA, and suppression of the flow signal in the target arterial region on 3D TOF-MRA was achieved. The BeamSAT pulse was set before the TOF sequence in each repetition time and was extended from 23.0 ms to 40.5 ms. To suppress only the target artery, a 30-mm diameter BeamSAT pulse was positioned to cover the target artery precisely.

ICA-selective MRA was performed by adjusting the insertion direction of the BeamSAT pulse to penetrate and suppress the flow signals of the three major vessels (contralateral ICA and both vertebral arteries) (Figure 1A). Only the target ICA flow signal remained, and an ICA-selective MRA scan was obtained (Figure 1B). VA-selective MRA was performed by adjusting the insertion direction of the BeamSAT pulse to penetrate and suppress the flow signals of the two major vessels (both ICAs), leaving only the VA flow signal.

**Figure 1.**
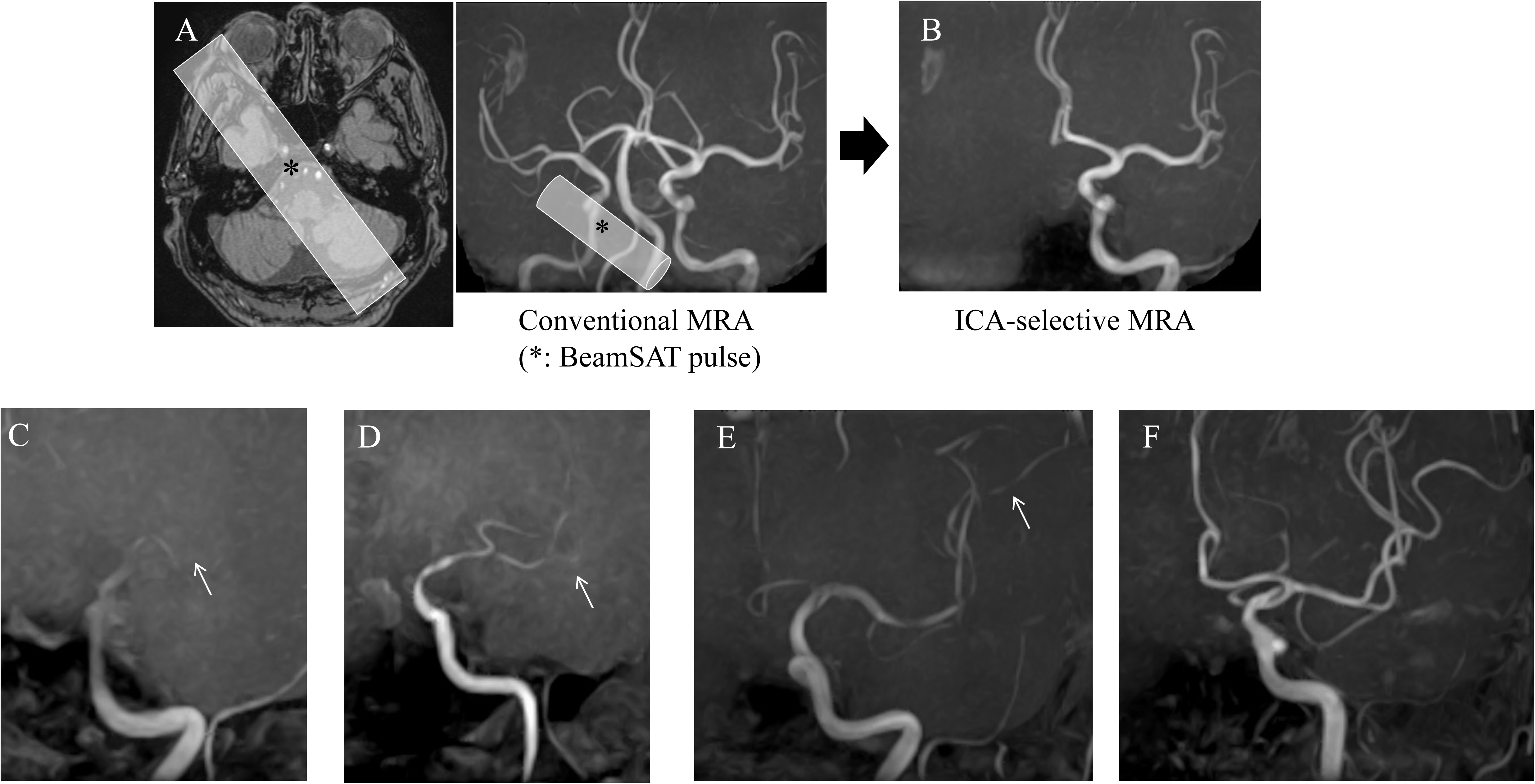
(A,B) The BeamSAT pulse is positioned to cover the unilateral petrous portion of the ICA and bilateral vertebral arteries in an axial TOF source image obtained with conventional MRA (A, left). By adding the BeamSAT pulse to the unilateral ICA and bilateral vertebral arteries on 3D TOF-MRA (A, right), we perform ICA-selective MRA (B). The asterisk indicates the BeamSAT pulse. (C-F) Degree of visualization of the ipsilateral MCA on brain MRA is graded as follows: (C) The M1 cannot be visualized along its course (grade 1, arrow). (D) One M2 branch can not be visualized along its course (grade 2, arrow). (E) One M3 branch cannot be visualized to the cortical surface (grade 3, arrow). (F) All M3 branches of the left MCA can be visualized to the cortical surface (grade 4).

### Image analysis

#### Preoperative assessment of cerebral perfusion status

Using BeamSAT MRI, we defined the crossflow through the circle of Willis based on the following criteria: cases that displayed a stenosed MCA being visible via either non-stenosed ICA-selective MRA or VA-selective MRA were deemed crossflow-positive. We evaluated variations in the depiction of the MCA between conventional MRA and ICA-selective MRA using the following specific criteria: the signal intensity of the MCA on both conventional and ICA-selective MRA was categorized into four distinct grades, termed the MRA-MCA grades, as previously reported.^16^ Grade 1 indicated that the M1 segment was not visible, as shown in Figure 1C. Grade 2 meant that at least one M2 branch was not visible, illustrated in Figure 1D. Grade 3 indicated that at least one M3 branch was not visible, as seen in Figure 1E. Lastly, Grade 4 specified that all M3 branches of the MCA were visible, depicted in Figure 1F. In a previous study, Hirooka et al. demonstrated the efficacy of predicting impaired CVR by categorizing MCA-MRA grades as 1 to 3 and grade 4 in conventional MRA.^16^ Consequently, we adopted a similar classification in our comparison of MCA grades 1 to 3 with grade 4 in MRA.

Cases exhibiting a grading discrepancy between conventional MRA and ICA-selective MRA were labeled as mismatch-positive. In all cases labeled as mismatch-positive, the grade on the ICA-selective MRA was lower than that on the conventional MRA. Patients with either crossflow-positive or mismatch-positive findings were classified into the developed collateral (DC) group, while all others were categorized into the undeveloped collateral (UC) group (Supplementary figure).

### SPECT

All patients were scanned preoperatively using a rotating dual-headed gamma camera with dynamic SPECT (ECAM GMS7700, Toshiba Medical, Tokyo, Japan). We used a dual-table autoradiographic (DTARG) method that was developed for use with diffusible tracers to quantify CBF at rest and after pharmacologic stress from a single session of dynamic scans with dual bolus administration of N-isopropyl-[123I]-p-iodoamphetamine.^17^ The SPECT scan was started immediately after the administration of IMP in both the rest and ACZ challenge scans. CBF was quantified using the QSPECT/DTARG method, which automatically and accurately corrects attenuated absorption and scattered radiation.^18^ We used the NEUROSTAT software program for the anatomical standardization of SPECT images. The regions of interest were automatically placed in both the cerebral and cerebellar hemispheres using the NEURO FLEXER template.^19^ The mean CBF in the resting state and after ACZ challenge was measured in each MCA territory (anterior and posterior). The CVR to ACZ challenge was calculated as follows: CVR (%)=[(ACZ challenge CBF–resting CBF)/resting CBF]×100. Following the CEA/CAS, only resting SPECT was conducted, with no ACZ challenge implemented. CBF in the anterior and posterior regions of the MCA was evaluated both pre- and post-operatively. All image analyses were carried out by two seasoned neurosurgeons, S.Y. and M.K., who were blinded to the patients’ clinical data.

### Evaluation of impaired CVR

When CVR of either the anterior or posterior MCA territory was less than 10%, which has been reported to indicate the risk of hyperperfusion in previous studies,^20,21^ it was rated as impaired CVR.

### Evaluation of postoperative HPS by SPECT

The following items were retrospectively investigated from the patients’ medical charts as symptoms of suspected HPS: multiple complaints of headache and nausea/vomiting, seizures, focal neurological deficits, and intracranial hemorrhage on imaging within one week after surgery.^3^

The anterior and posterior MCA asymmetry index values were calculated as the ratio of the stenosed side to the non-stenosed side within the respective MCA territories.^20^ The CBF pre/post ratio, representing the ratio of postoperative to preoperative CBF, was calculated by comparing the postoperative MCA asymmetry index with the preoperative MCA asymmetry index. The larger value between the anterior and posterior CBF pre/post ratio was denoted as the MCA CBF pre/post ratio. Patients with a high MCA CBF pre/post ratio were interpreted as having a substantial increase in CBF following the intervention.

### Statistical analysis

Descriptive statistics are presented as mean±standard deviation and were compared using the Welch two-sample t-test. The proportion of patients with each parameter was compared using the Fisher exact test. The significance of differences between more than two groups was determined using the Tukey–Kramer test.

Regarding the multivariate analysis for impaired CVR, being in the DC group, being symptomatic, and having high PSV (>200cm/s) were selected as the independent predictor variables, according to the clinical literature and expert opinion. The predictive accuracy of the logistic regression model was quantified by using the area under curve (AUC) of the receiver operating characteristic (ROC) curve. The confidence intervals of the AUC were calculated using the bootstrap percentile method. Partial AUC (pAUC) was computed in the portion of the ROC space where the true positive rate (sensitivity) was greater than a given threshold.

Statistical analyses were performed using open-source software (R4.3.0; R Foundation for Statistical Computing, Vienna, Austria; http://www.r-project.org/). Differences were considered statistically significant at P<0.05. The ROC curve and pAUC were computed using the pROC package of R.^22^

## Results

### Patient characteristics

During the time of the study, 48 patients (27 CEA and 21 CAS) fulfilled the inclusion criteria. Forty-one patients were male and 7 were female. The mean age was 76.3 + 6.13 years. The overall average of the degree of ICA stenosis was 76.3 + 11.9%, according to the NASCET criteria. Twenty-four patients were symptomatic, and 24 were asymptomatic.

In our cohort of 48 patients, conventional MRA identified 8 patients with an MCA-MRA grade 3 and 40 with an MCA-MRA grade 4. In contrast, ICA-selective MRA revealed 31, 9, 5, and 3 patients with MCA-MRA grades 4, 3, 2, and 1, respectively. Of the 40 patients identified with an MCA-MRA grade 4 by conventional MRA, 6 were graded as 3 on ICA-selective MRA, 1 was graded as 2 on ICA-selective MRA, and 2 were graded as 1 on ICA-selective MRA, indicating a mismatch-positive status. The remaining 31 patients’ condition did not change from grade 4. Of the 8 patients identified as MCA-MRA grade 3 by conventional MRA, 4 were graded as 2 on ICA-selective MRA, and 1 was graded as 1 on ICA-selective MRA, indicating a mismatch-positive status. The remaining 3 patients’ condition did not change from grade 3. There was an MCA-MRA grade mismatch between the conventional and ICA-selective MRA techniques in 14 patients. In all cases labeled as mismatch-positive, the grade on the ICA-selective MRA was lower than that on the conventional MRA.

Furthermore, crossflow from the contralateral ICA or VA was observed in 10 patients, and of these, 5 exhibited the aforementioned mismatch. Crossflow was observed in 5 of 34 patients with MCA-MRA grade match. Therefore, on the basis of the presence or absence of collateral vessels, 19 patients were categorized into the DC group, whereas the remaining 29 were categorized into the UC group (Supplementary figure).

Table 1 summarizes the baseline characteristics of the 48 patients analyzed in this study. There were significant differences in baseline characteristics between the DC and UC groups. Age, degree of ICA stenosis, PSV, and frequency of hyperlipidemia were significantly higher in the DC group than in the UC group (79.0±4.6 versus [vs.] 74.6±6.42, P=0.0081; 81.9±7.64% vs. 72.1±13.1%, P=0.045; 344.5±123.1 cm/s vs. 245.4±113.7 cm/s, P=0.009; 8/19 vs. 22/29, P=0.032, respectively). The preoperative ipsilateral CVR was significantly lower in the DC group than in the UC group (18.0±20.0% versus [vs.] 48.3±19.5%; P<0.0001; Figure 2A). Preoperative frequency of impaired CVR was significantly higher in the DC group than in the UC group (8/19 vs. 1/29, P=0.004).

**Table. 1.** Specific characteristics of patients between the developed collateral (DC) group and undeveloped collateral (UC) group.

| Characteristic | DC (n=19) | UC (n=29) | P Value |
| --- | --- | --- | --- |
| Sex Male | 17 (89.4%) | 23 (79.3%) | 0.45 |
| Age in yrs | 79.0 ± 4.6 | 74.5 ± 6.4 | 0.008 |
| Rt. ICS | 9 (47.4%) | 13 (44.8%) | 1 |
| CEA | 10 (52.6%) | 17 (58.6%) | 0.77 |
| Symptomatic | 9 (47.4%) | 15 (51.7%) | 1 |
| Degree of stenosis, % | 81.9 ± 7.6 | 72.1 ± 13.1 | 0.045 |
| PSV, cm/s | 344.5±123.1 | 245.4±113.7 | 0.009 |
| Hypertension | 12 (63.2%) | 25 (86.2%) | 0.085 |
| Hyperlipidemia | 8 (42%) | 22 (76%) | 0.032 |
| Diabetes mellitus | 4 (21.1%) | 13 (44.8%) | 0.12 |
| Ischemic heart disease | 6 (31.6%) | 9 (31.0%) | 1 |
| Smoking | 12 (63.2%) | 13 (44.8%) | 0.24 |
| COPD | 3 (15.8%) | 2 (6.9%) | 0.37 |
| Ipsilateral CVR, % | 18.0±20.0 | 48.3±19.5 | <0.0001 |
| Impaired CVR (CVR<10%) | 7 (36.8%) | 1 (3.4%) | 0.004 |
| Symptoms suggestive of HPS | 8 (42%) | 1 (3.4%) | 0.001 |
| MCA CBF pre/post ratio | 1.092 | 1.026 | 0.005 |

Values are reported as mean ± SD or number (%)
Rt, right; ICS, internal carotid artery stenosis; CEA, carotid endarterectomy; PSV, peak systolic velocity; COPD, chronic obstructive pulmonary disease; CVR, Cerebrovascular Reactivity; HPS, Hyperperfusion syndrome; MCA, middle cerebral artery; CBF, cerebral blood flow

**Figure 2.**
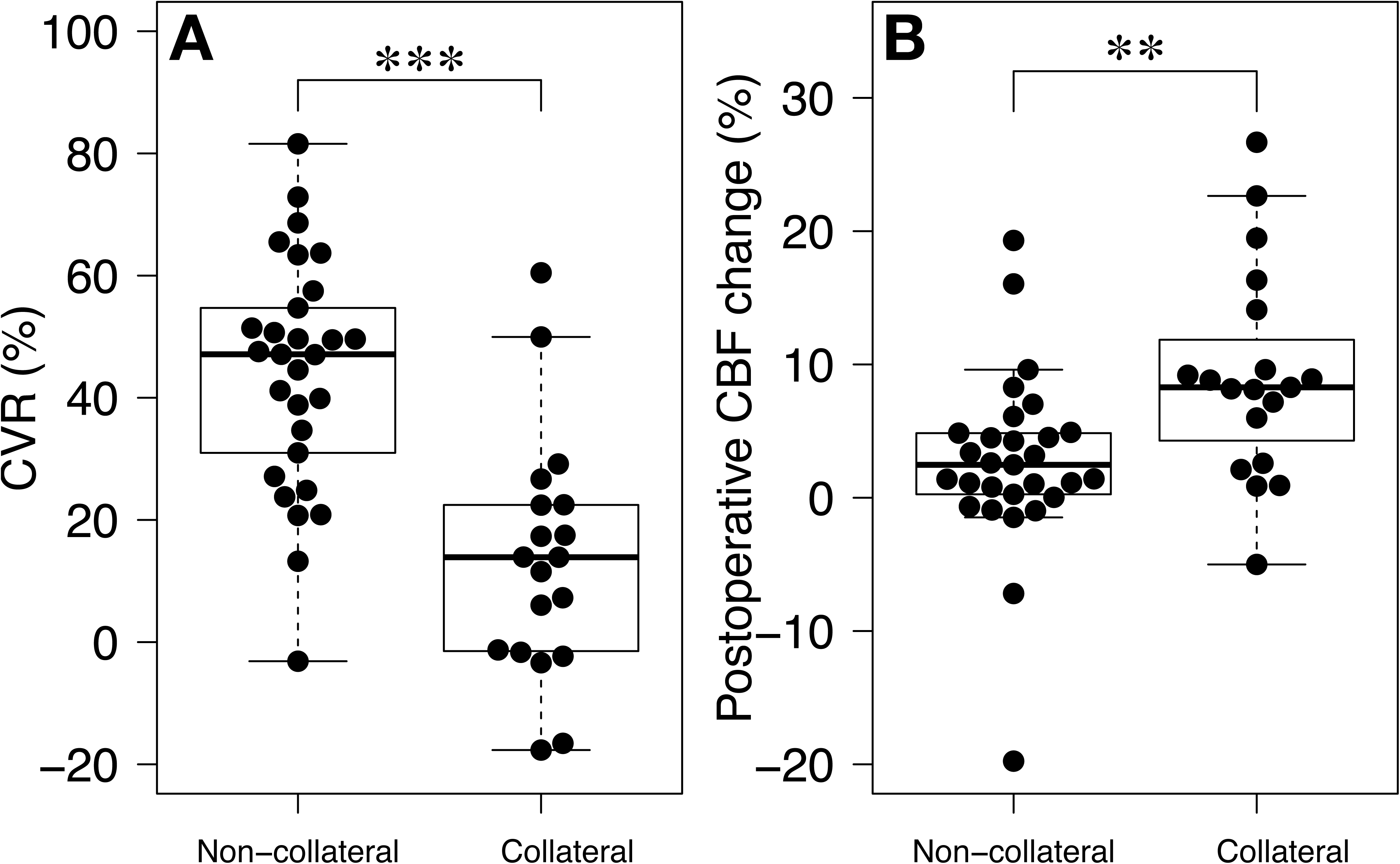
(A) Box-and-whisker plots of preoperative CVR on the stenotic side in the developed collateral (DC) group and the undeveloped collateral (UC) group, showing significantly lower in the DC group than in the UC group. Triple asterisks indicate P < 0. 001. (B) : Box-and-whisker plots of postoperative CBF changes in the ipsilateral MCA region in the UC and DC groups, showing significantly higher values in the DC group than in the UC group. Double asterisks indicate P < 0. 01. Median values are represented by thick horizontal lines within the boxes, with the first and third quartiles forming the box boundaries. Whiskers extend to the most extreme data points not considered outliers, no more than 1.5 times the interquartile range from the box.

### Univariate analysis and multivariate analyses

The following items were adopted as confounders in the logistic regression model for multivariate analysis (BeamSAT model): the DC group, high PSV (>200cm/s), and symptoms. The odds ratios for these variables in the univariate and multivariate analyses are listed in Table 2A. The DC group (odds ratio [OR] = 17.7, [95%CI: 1.83 to 171]; P = 0.013) was significantly associated with the development of impaired CVR. The discriminative value of this logistic regression model, as expressed by the AUC, was 0.858 (95%CI: 0.754-0.961), which indicated some utility in predicting the response of individual patients. The sensitivity and specificity of this logistic regression model for diagnosing impaired CVR were 88.9% and 71.8%, respectively. In contrast, the logistic regression model with the MCA-MRA grades (grade 1, 2, and 3 vs. 4), high PSV (>200 cm/s), and symptoms as predictor variables (conventional model) did not show a significant association with the outcomes (Table 2B). The full AUC of the BeamSAT model (0.858 [95%CI: 0.754-0.961]) was larger than that of the conventional model (0.698 [95%CI: 0.497-0.899]), although the difference in the full AUCs between the two models was not statistically significant (P = 0.113) (Figure 3A and B). Since we were interested in a clinical test with a high sensitivity for screening, we focused on the pAUC between 0.6 and 1 sensitivity. The estimates of the pAUC were computed by integration over the ranges starting at 1 and ending at 0.9, 0.8, 0.7, 0.6, and 0. Table 3 summarizes the data of pAUCs. The differences in the full AUCs was not statistically significant (P = 0.113), while the difference in the pAUCs over the range (1-0.6) of sensitivity was statistically significant (P = 0.04) (Figure 3C).

**Table. 2.** Univariate and multivariate analyses examining factors associated with impaired CVR (CVR <10%)

A: BeamSAT model
| Variables | Univariate |  |  | Multivariate |  |  |
| --- | --- | --- | --- | --- | --- | --- |
|  | OR | 95% CI | P value | OR | 95% CI | P value |
| PSV (>200cm/s) | 3.56 | 0.40-31.7 | 0.25 | 2.46 | 0.17-34.5 | 0.51 |
| Symptomatic | 2.11 | 0.46-9.64 | 0.34 | 3.15 | 0.50-19.7 | 0.22 |
| DC group<br>(vs. UC group) | 20.4 | 2.27-183 | 0.007 | 17.7 | 1.83-171 | 0.013* |

B. Conventional model
| Variables | Univariate |  |  | Multivariate |  |  |
| --- | --- | --- | --- | --- | --- | --- |
|  | OR | 95% CI | P value | OR | 95% CI | P value |
| PSV (>200cm/s) | 3.56 | 0.40-31.7 | 0.25 | 6.71 | 0.64-70.0 | 0.11 |
| Symptomatic | 2.11 | 0.46-9.64 | 0.34 | 4.11 | 0.74-22.9 | 0.11 |
| MCA-MRA grade 1/2/3<br>(vs. grade 4) | 1.57 | 0.26-9.47 | 0.45 | 2.31 | 0.32-16.5 | 0.40 |

CVR, Cerebrovascular reactivity; PSV, peak systolic velocity; OR, odds ratio; CI, confidence interval; DC, developed collateral; UC, undeveloped collateral; MCA, middle cerebral artery; MRA. Magnetic resonance angiography; \*: statistically significant

**Table. 3.**
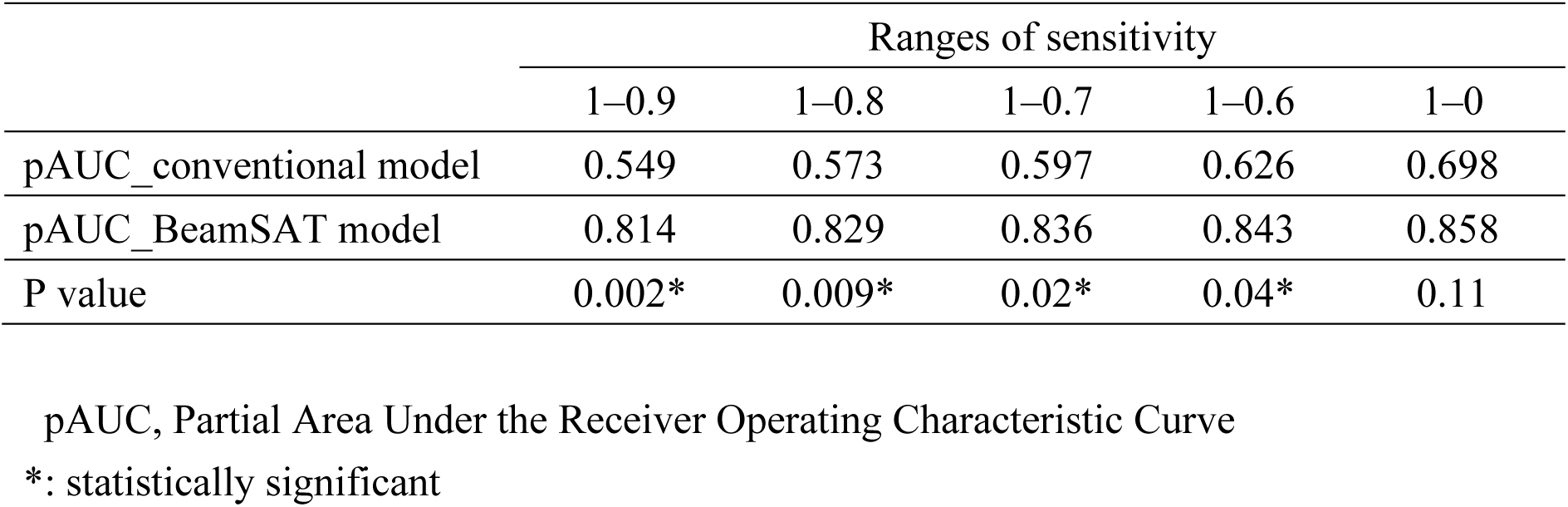
The summarized data of estimated pAUC.

**Figure 3.**
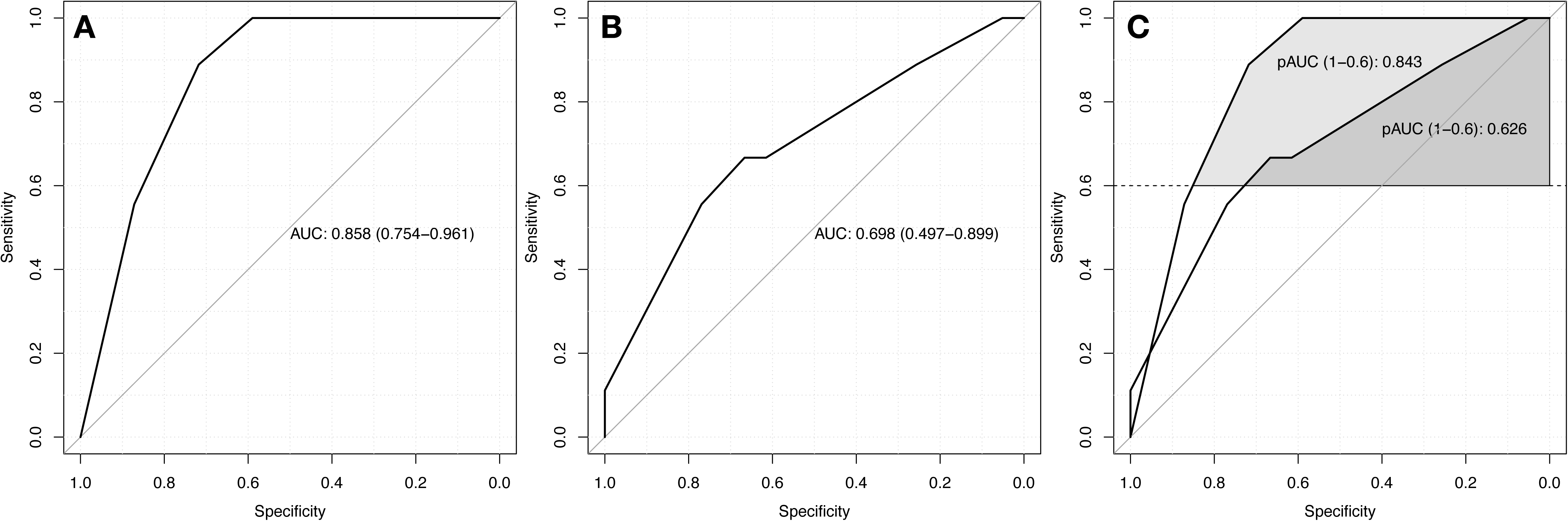
ROC curves of parameters for the prediction of impaired CVR. (A) : ROC curve of the BeamSAT model for impaired CVR. (B) : ROC curve of the conventional model for impaired CVR. The full AUC of the curve is printed in the middle of the plot, with 95% confidence interval in A and B. (C) : ROC curves of the BeamSAT model and conventional model. The partial AUC (pAUC) of both curves is printed in the middle of the plot, with 95% confidence interval. See text for details.

### Postoperative changes in CBF and symptoms suggestive of HPS

Postoperatively, the incidence of symptoms suggestive of HPS was significantly higher in the DC group than in the UC group (42% [8/19] vs. 3% [1/29], P=0.001). The MCA CBF pre/post ratio was significantly elevated in the DC group (1.092) compared with the UC group (1.026) (P=0.005) (Figure. 2B).

## Discussion

In this study, we used a minimally invasive method, BeamSAT MRI, to evaluate impaired CVR, which is considered to be a high-risk factor for future cerebral infarction and postoperative HPS.

The current results demonstrated that the DC group was significantly associated with impaired CVR. The value of the AUC (0.858) of the current logistic regression model (BeamSAT model) indicated some utility in prediction. In the prevention of HPS, it is important to identify patients with impaired CVR, i.e. high sensitivity is the region of interest. Full AUCs may not show significant differences between different models, even though they different in the regions of interest. In the current study, full AUCs did not show a significant difference between the two groups, although the full AUC was larger in the BeamSAT model than in the conventional model. However, the pAUC at the cut-off value of more than 0.6 sensitivity showed a considerable difference in performance between the BeamSAT model and conventional models, showing that the BeamSAT model performed much better (Table 3).

Interestingly, patients classified in the DC group on the preoperative BeamSAT MRI showed a significant increase in the CBF ratio between the preoperative and postoperative assessments compared with the UC group. Moreover, the DC group showed a significantly higher incidence of suggestive HPS symptoms than the UC group. Considering these findings, BeamSAT MRI can be a useful and minimally invasive tool for the prediction of impaired CVR and postoperative HPS.

Sebok and Sobczyk et al. observed a significant correlation between CVR and collateral development.^9,10^ The pattern of collateral blood flow resulting from carotid artery stenosis can be categorized into primary pathways, such as those through the circle of Willis from the contralateral ICA and VA, and secondary pathways, including routes via the external carotid artery, ophthalmic artery, and leptomeningeal vessels.^23^ In our study, collateral flow evaluations were primarily focused on routes other than leptomeningeal anastomoses. Therefore, extra vigilance may be required in the management of patients with an inadequate circle of Willis and severe stenosis.

In our current analysis, BeamSAT MRI findings that indicated the existence of collateral flow were strongly linked to impaired CVR. This underscores the concept that as CVR decreases, collateral flow through the circle of Willis is augmented to preserve CBF. Lan et al. confirmed this finding by showing that the ratios of signal intensity on TOF-MRA correlate with cerebral perfusion, as measured by SPECT, in adults with intracranial carotid stenosis.^24^ Our analysis revealed that the MCA signal intensity varied between conventional MRA and ICA-selective MRA. In some cases, the MCA appeared to be more subdued on the ICA-selective MRA. These differences led us to infer the existence of collateral flow. Various modalities, including PET, SPECT, perfusion CT, and ASL, have been explored to assess hemodynamic insufficiency. It is worth noting that SPECT and PET require radiation exposure and the administration of radioactive tracers. There have also been instances of adverse reactions such as pulmonary edema following ACZ use during CVR evaluation using SPECT.^25^ Sasaki et al. identified that a stage marked by moderately reduced CBF, extended mean transit time, and augmented CBV on perfusion CT was indicative of misery perfusion.^26^ However, it is important to acknowledge the inherent risks of radiation exposure with perfusion CT in addition to the requirement for an iodinated contrast agent.

Recently, advancements in MRI methodologies, such as ASL, have emerged as viable alternatives for CVR measurement.^27^ However, Kimura et al. reported that ASL tends to underestimate the CBF in patients with chronic major artery stenosis or occlusion.^28^ Taken together with these previous studies, the current study suggests that BeamSAT is a non-invasicve and accessible modality for investigating cerebral hemodynamics.

HPS is a complex clinical phenomenon that can occur after revascularization procedures such as CEA and CAS.^3,29^ It has been reported that the mortality rate due to HPS is up to 17% in patients with ICH.^30^ Treatment for HPS includes lowering the blood pressure and bed rest, but there is no definitive cure; thus, prevention is the key. Preoperative impairment of cerebrovascular autoregulation is thought to be a risk factor for HPS, requiring careful treatment and postoperative management, especially if impaired CVR was confirmed preoperatively.^3^ In this study, the DC group patients, who were suspected of impaired CVR, had a significantly higher risk of symptoms suggestive of HPS and a higher rate of significant CBF increase after surgery than the UC group. These findings suggest that BeamSAT MRI may be used to predict the possibility of HPS.

Our study has several limitations. First, care must be taken in patients who anatomically do not have collateral blood circulation channels such as the Acom or Pcom, or who have stenosis in the contralateral ICA or VA. Second, our MRI technique was unable to evaluate the collateral flow through the cortical leptomeningeal artery. Third, BeamSAT MRI is available only for the 1.5T ECHELON Smart Plus (FUJIFILM Healthcare Corporation, Tokyo, Japan) model. Fourth, the scope of our study was limited, and only a small patient cohort was examined. Lastly, given that these are retrospective data drawn from a modest sample size, there is potential for overfitting, meaning that our findings might be overly tailored to our specific dataset. Further research on a broader scale is required to validate our conclusions.

## Conclusions

In patients with carotid artery stenosis, BeamSAT MRI demonstrated high capability for predicting impaired CVR, which is a major risk factor for postoperative HPS, with minimal invasiveness. Patients who demonstrate collateral flow on BeamSAT MRI and have different MCA-MRA grades between ICA-selective and conventional MRA are at risk of impaired CVR. Consequently, BeamSAT MRI may be a valuable non-invasive tool for assessments in individuals with carotid artery stenosis.

## Data Availability

The data that support the findings of this study are available from the corresponding author, but restrictions apply to the availability of these data, which were used under license for the current study, and so are not publicly available. Data are however available from the authors upon reasonable request and with permission of the corresponding author.

## Author Contributions

## Acknowledgments

None.

## Sources of Funding

This study received no specific grants from funding agencies in the public, commercial, or non-profit sectors.

## Disclosure

None.

**Supplementary figure.** In our 48-patient cohort, MCA-MRA grade discrepancies between conventional and ICA-selective MRA techniques were observed in 14 patients, while grade concordance was noted in 34 patients. Crossflow was present in 5 of the 34 patients with matching MCA-MRA grades. On the basis of the presence or absence of collateral vessels, 19 patients were categorized into the developed collateral (DC) group, whereas the remaining 29 patients were categorized into the undeveloped collateral (UC) group.

## Notes

### Competing Interest Statement

The authors have declared no competing interest.

### Author Declarations

This study received approval from the Institutional Review Board (IRB) of Kobe University Hospital. The approval number for this study is #170024. In compliance with the ethical guidelines and regulations set by the IRB, written informed consent was obtained from all participants involved in this study. This process ensured that all participants were fully informed about the nature of the research, its objectives, and the use of the data collected, and their participation was voluntary.

